# FAMILY SUPPORT-BASED PSYCHOEDUCATION IN CARING SCHIZOPHRENIC PATIENTS: A LITERATURE REVIEW

**DOI:** 10.1101/2022.01.11.22268923

**Authors:** Dwi Indah Iswanti, Moses Glorino Rumambo Pandin

## Abstract

**Background:** The burden of family in caring a schizophrenia impacts on psychological shock and emotional burden; lack of disease knowledge and care skills; poor patient medication adherence; the difficulty of getting along with patients; conflict in the family or at work; financial burden; and need adequate social support.

**Aim:** This study discussed how family support takes care of people with schizophrenia with a psychoeducational approach in mental health service settings.

**Method:** A non-systematic literature review was carried out using the PICO (Population, Intervention, Comparison, Outcome) framework utilizing Scopus, CINAHL, and ProQuest databases in the last 5 years, searching with keywords “Family Support AND Schizophrenia OR Family Psychoeducation”.

**Results:** Families with schizophrenia experience the most potent stressors on the burden of care borne, in addition to stigma and the frequency of relapse of sufferers. Community care providers can involve the family in various ways, such as providing psychoeducation, supporting the family’s physical, emotional and social needs, and behavioral family assessment or family therapy.

**Conclusion:** Mental health services must develop and imply family psychoeducation training.

## INTRODUCTION

Schizophrenia is a chronic disease that requires long-term care. Barriers that families often encounter in caring for schizophrenia include isolation of patients from daily life, incomplete recovery, lack of support by the mental health care system, and stigmatization.^1^ The barriers make burden for the family causing psychological shock and emotional burden; lack of disease knowledge and care skills; poor medication adherence from the patient; difficulty to get along with patients; conflict in the family or at work; financial burden; and require sufficient social support.^2^ This makes the family experience stress triggered by the limitations of the adequacy of the family structure and function, which will inhibit the family from using other resources and create family stress.^3^

The prevalence of schizophrenia in the world globally is more than 20 million people, there are 69%, and even more in the world, people with schizophrenia do not get appropriate services. 90% of people with schizophrenia undergo treatment and are in developing countries where the community’s income level is in the middle to low range. The problem is that families with schizophrenia have less access to mental health services, then, schizophrenia often requires treatment compared to other chronic diseases.^4^ Mental disorders are a significant public health problem and rank among the top 10 causes of the global disease burden.^5^ The prevalence of Schizophrenia in Indonesia was 7 cases per mile in 2018 (an increase of 0.18%).^6^

Families caring for schizophrenia will experience a parenting burden, where primary family caregivers experience a burden from mild to moderate levels and have a poor quality of life. The caregiver’s age and the condition of not having a job add to caregivers’ burden for people with schizophrenia. In addition, the family burden is also increased by difficulties in caring for sufferers with severe psychiatric symptoms, decreased cohabitation, and low family coping strategies that lead to poor quality of life.^5^ Family-focused interventions for institutionalized caregivers of people with schizophrenia that include psychological support and peer support groups are recommended to improve mutuality and family coping strategies, reduce caregiver burden, and improve caregiver quality of life..^5^ Thus, this literature review discusses family support through psychoeducation to lessen the burden of parenting and increase the capacity of families in treating schizophrenia.

## METHOD

Briefly, to obtain the evidence regarding the psychoeducation used by family as a support for caring the schizophrenia patients, we first identified published, peer-reviewed research articles using a non-systematic search across several databases. After processing eligible studies, we identified and obtained primary studies from these published research papers to generate the compendium of papers about psychoeducation used by family as a support for caring the schizophrenia patients.We developed a narrative summary and a thematic synthesis to synthesize the quantitative and qualitative data, respectively. We conducted third synthesis to combine the previous two syntheses. No protocol existed for this literature review. This research did not involve participation of human subjects, thus research ethics board review was not required, nor was informed consent.

### Search strategy and inclusion criteria for literature reviews

The literature review was performed using the PICO Literature Review (Table 1) framework within ScienceDirect, ProQuest, SAGE, DOAJ (Directory of Open Access Journals)databases in the last 5 years, with keywords Family Support AND Schizophrenia OR Family Psychoeducation. In addition, to get many references, researchers also use synonyms in searching literature such as mental illness and mental health. Reviews were also carried out on abstracts and references from articles collected.

**Table 1.** Description of PICO Literature Review

|  |  |  |
| --- | --- | --- |
| P | Patient, Population, Problem | Schizophrenia, Mental illness |
| I | Intervention | Family support, Family Psychoeducation |
| C | Comparison Intervention (if appropriate) | NA |
| O | Outcome to Measure or Achieve | Treatment in schizophrenia |
| S | Study design | Quantitative, Qualitative and Random Control Trial |

After getting 20 eligible articles, the primary substance is identified and generalized as a psychoeducation-based family support theme.

Titles and abstract of manuscript were assessed for relevance to the topic using the following inclusion criteria:

1. The research design used quantitative, qualitative and Random Control Trial (RCT) published from 2017-2021 in English
2. Research respondents diagnosed with schizophrenia
3. The intervention approach used were in the form of family support and psychoeducation
4. Family support for treating schizophrenic patients with a psychoeducational approach was described

If relevance was not apparent based on the initial review, the full article was read to determine whether it should be included. After the initial group of articles was collected, 59 full-text articles were read to ensure final inclusion in the review. 20 articles met the inclusion criteria. Data were extracted into the JCR to review the study characteristics and the level of evidence level for each article. There were 5 articles as the evidence base of this study, 15 as professional standards, and 5 (recommendations) were not included in this study, because the purpose of this review was to investigate the original empirical studies. We focus on articles that highlight family support providing care for schizophrenia with a psychoeducational approach in primary mental health care.

### Study Selection, Data extraction and Management

From each full-text, we collected the data about the publication year, the databases searched, the study population, and the environment modifications. The authors independently screened titles, abstract, and full texts of articles. The authors retrieved full-text versions of potential articles and determined final inclusion in the review on the basis of relevance to the question, study quality, level of evidence, and inclusion and exclusion criteria. Disagreements were resolved by discussion or by consulting another author if necessary.

Each retained article was appraised and key information extracted to an evidence table that provide a summary of the methods and findings of the article. Supplemental table summarizes the characteristics, including methodology, environmental modification assessed, result, and recommendation.

### Risk of bias assessment and grading the quality of evidence

The risk of bias of each study was not evaluated. The overall quality of evidence was not assessed. Results regarding the risk of bias and grading the quality were resolved through discussion.

### Outcome measures

We focused on articles that considered psychoeducation as family support in caring patients with schizophrenia.

## RESULT AND DISCUSION

### Study selection

A total of 59 citations and abstracts were screened during the initial search. 29 records were excluded in the abstract review and 20 were assessed in full-text and following full-text review and ultimately, then eight studies were included in the final analysis (Figure 1).

**Figure 1.**
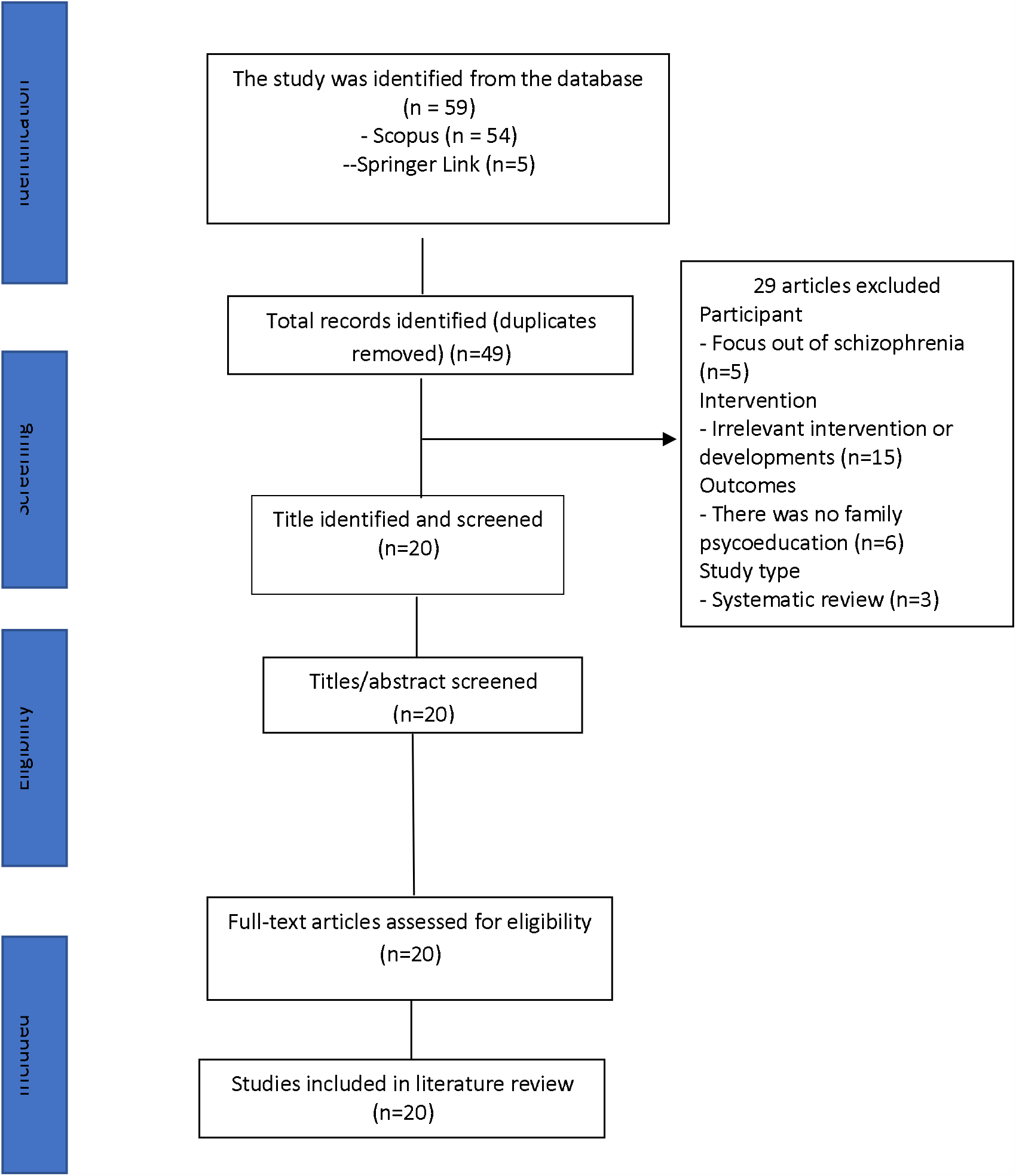
PRISMA Flow Diagram of Included Studies

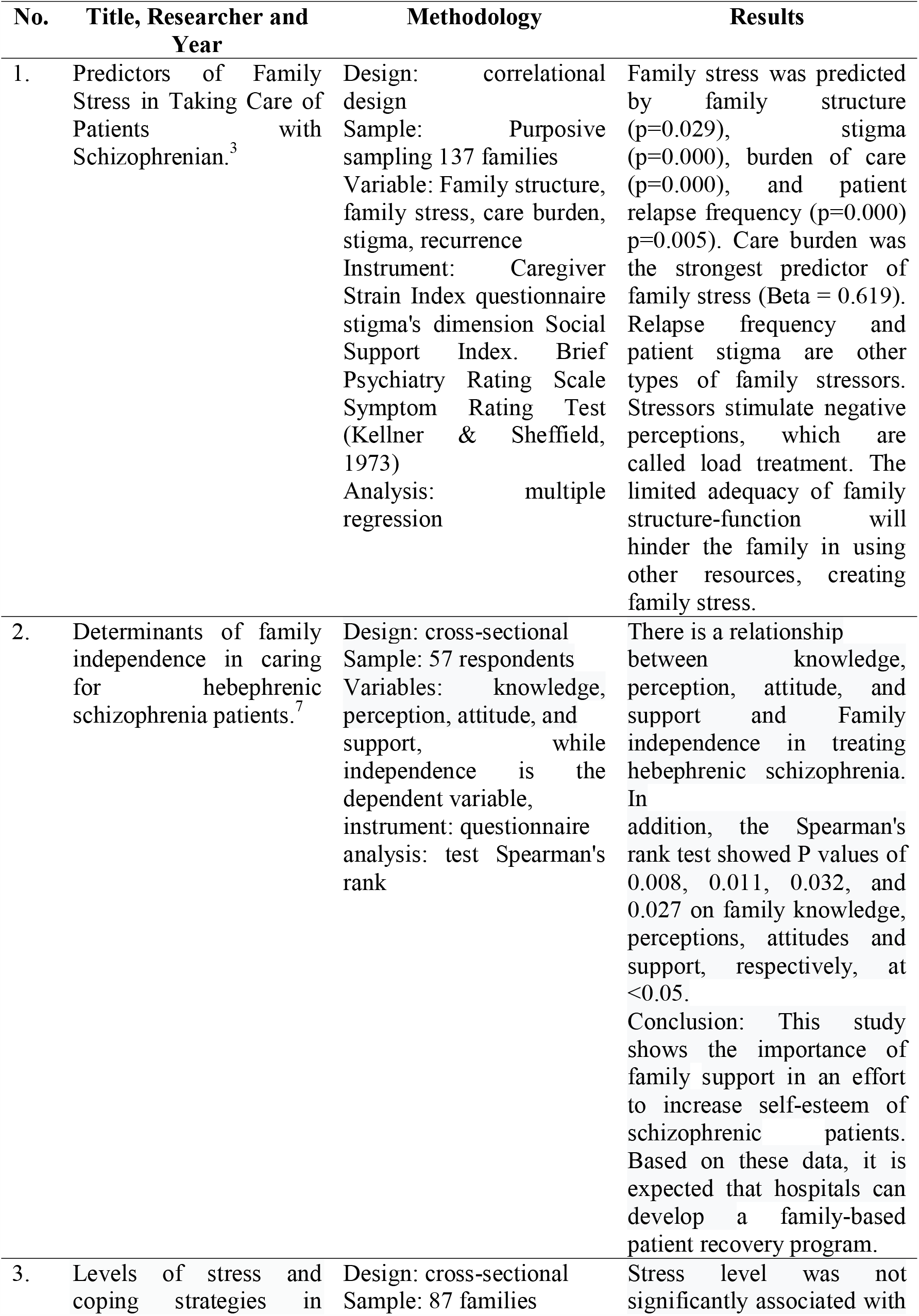
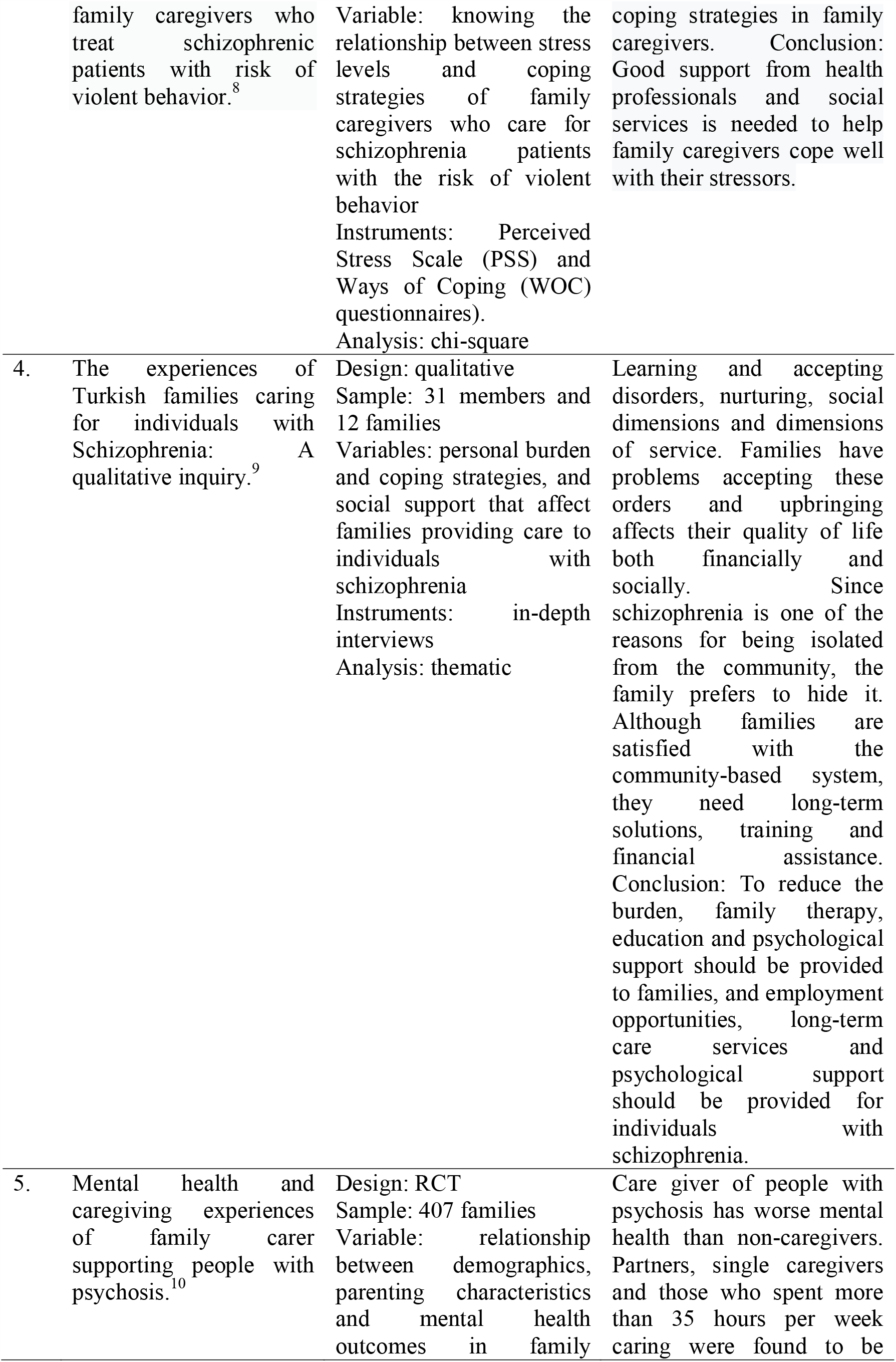
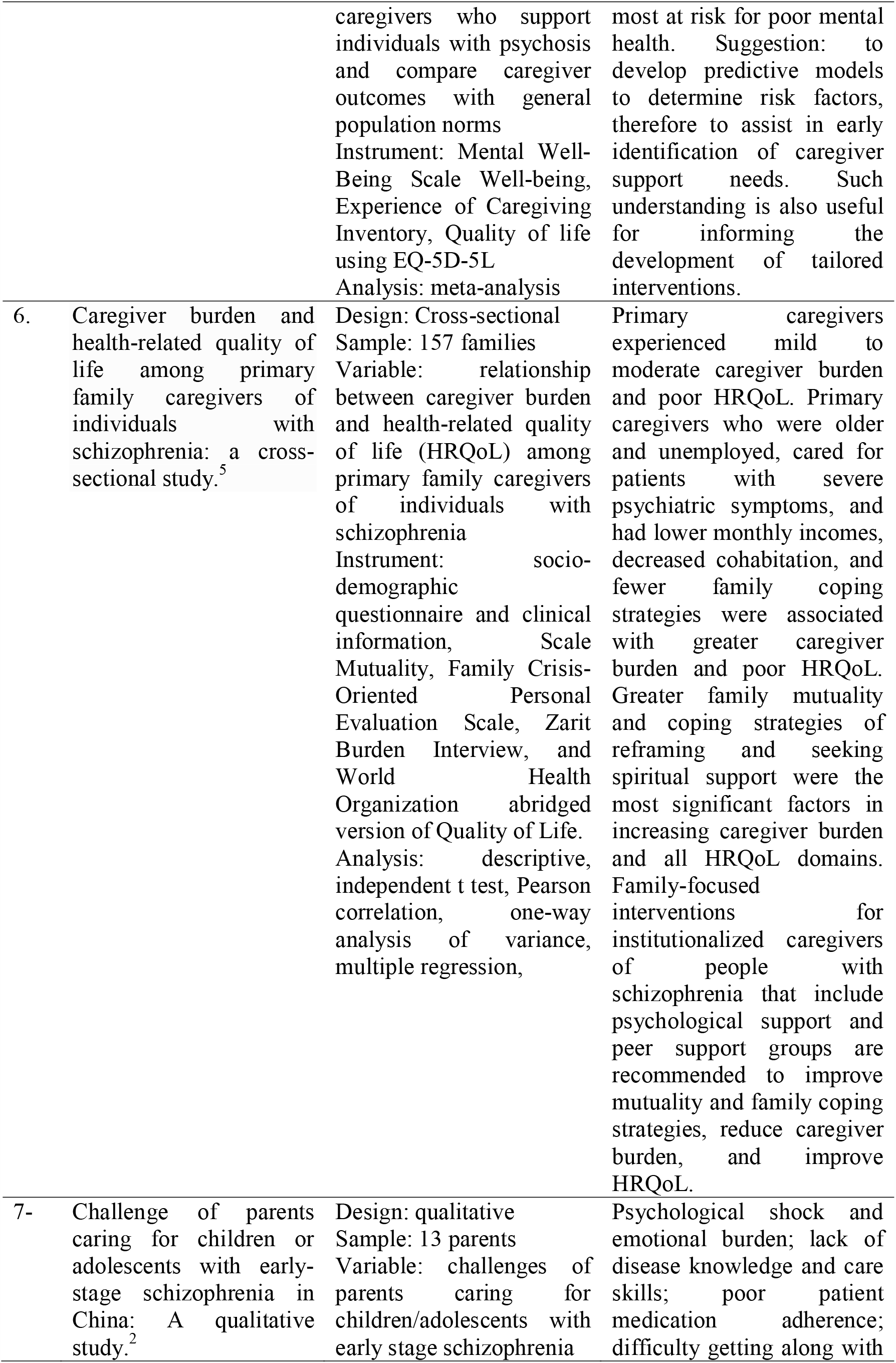
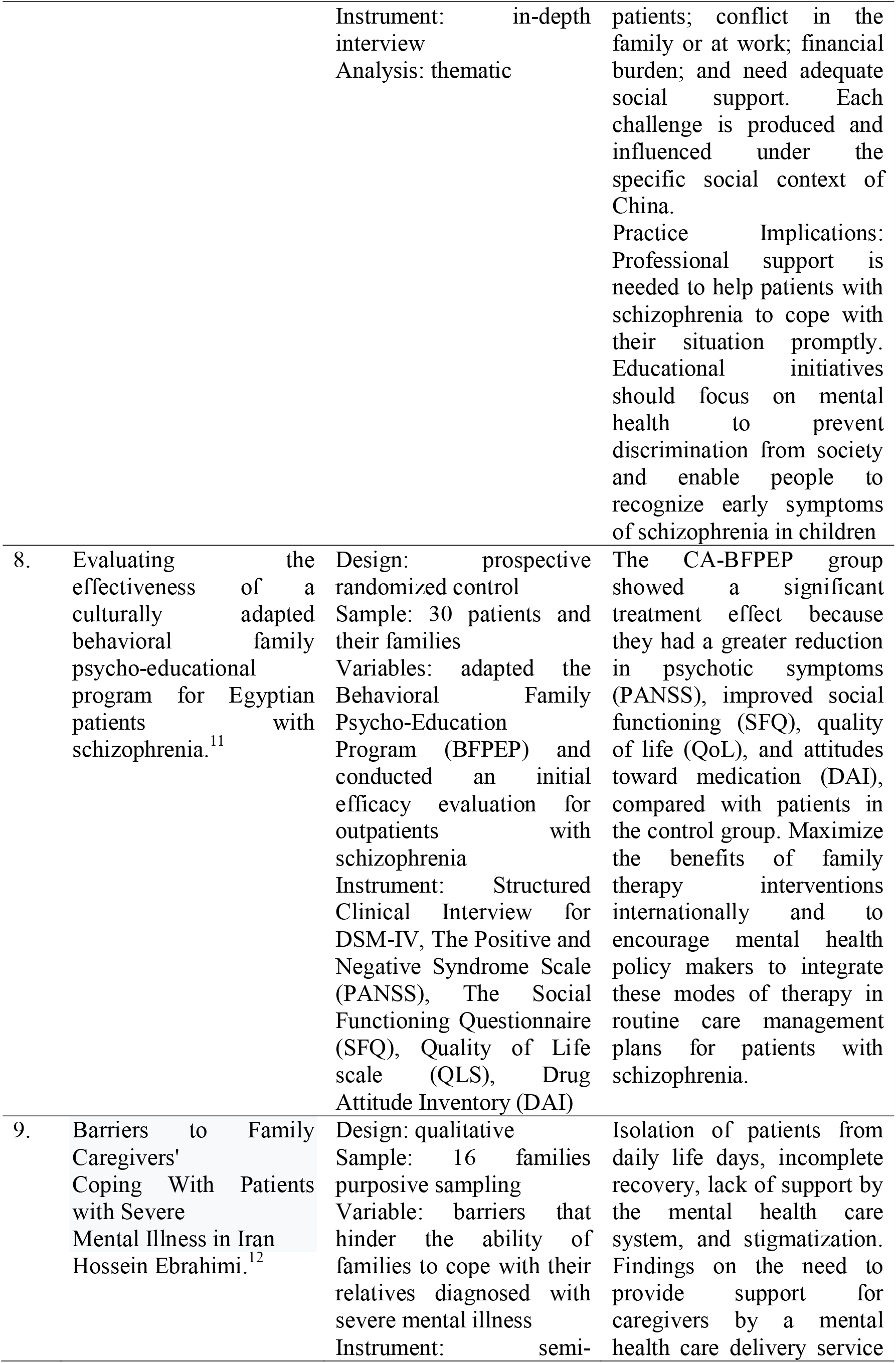
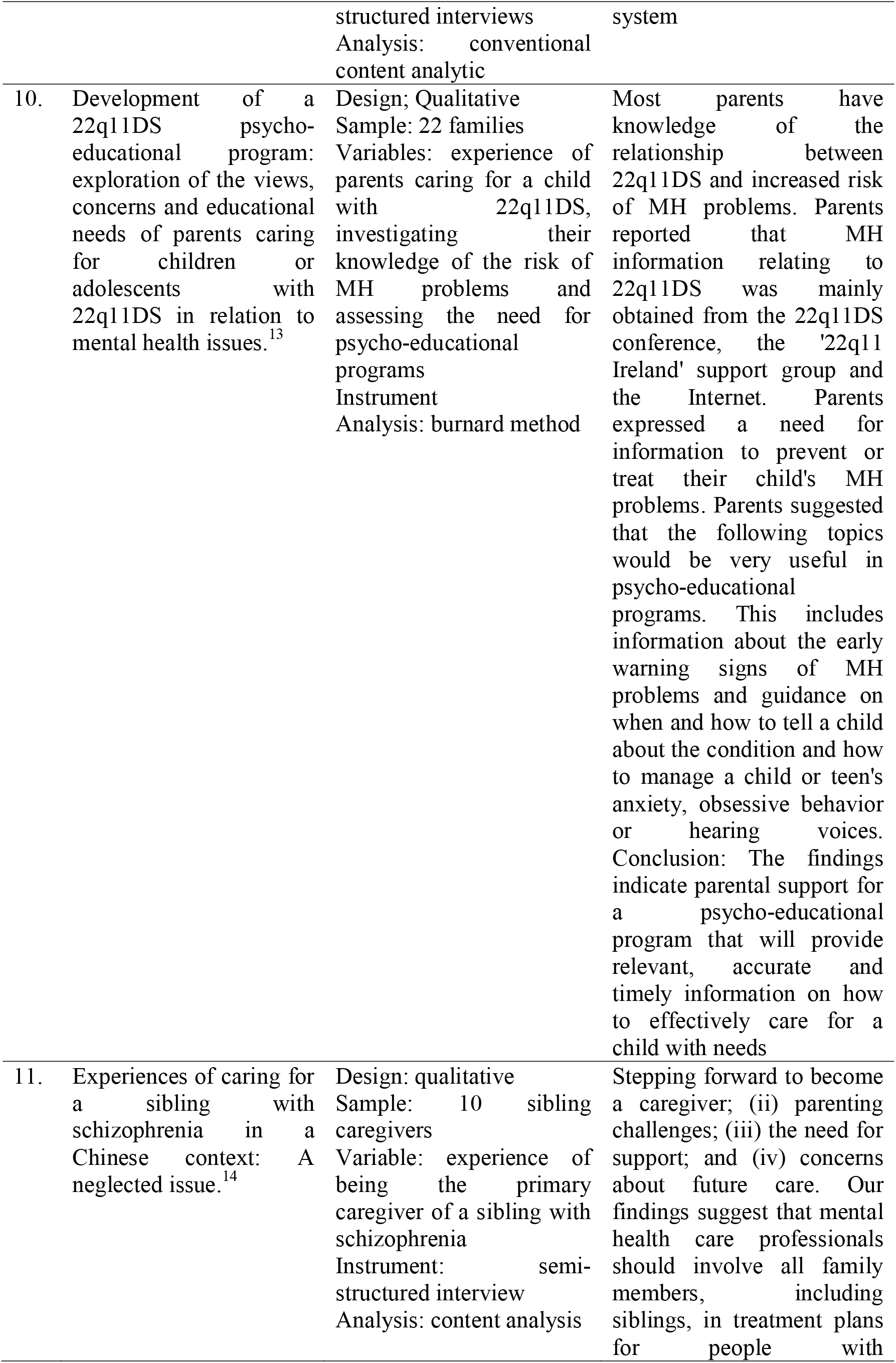
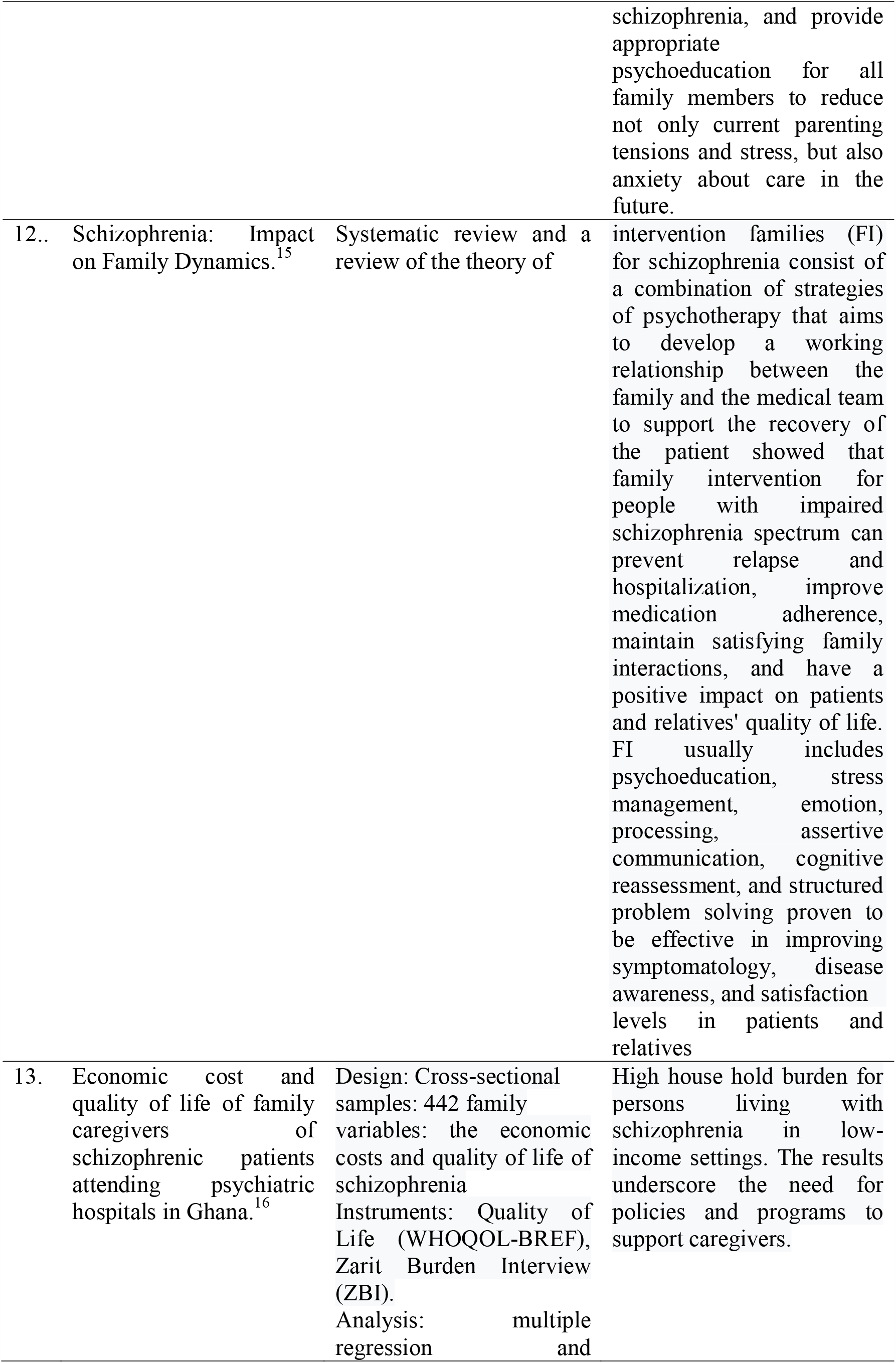
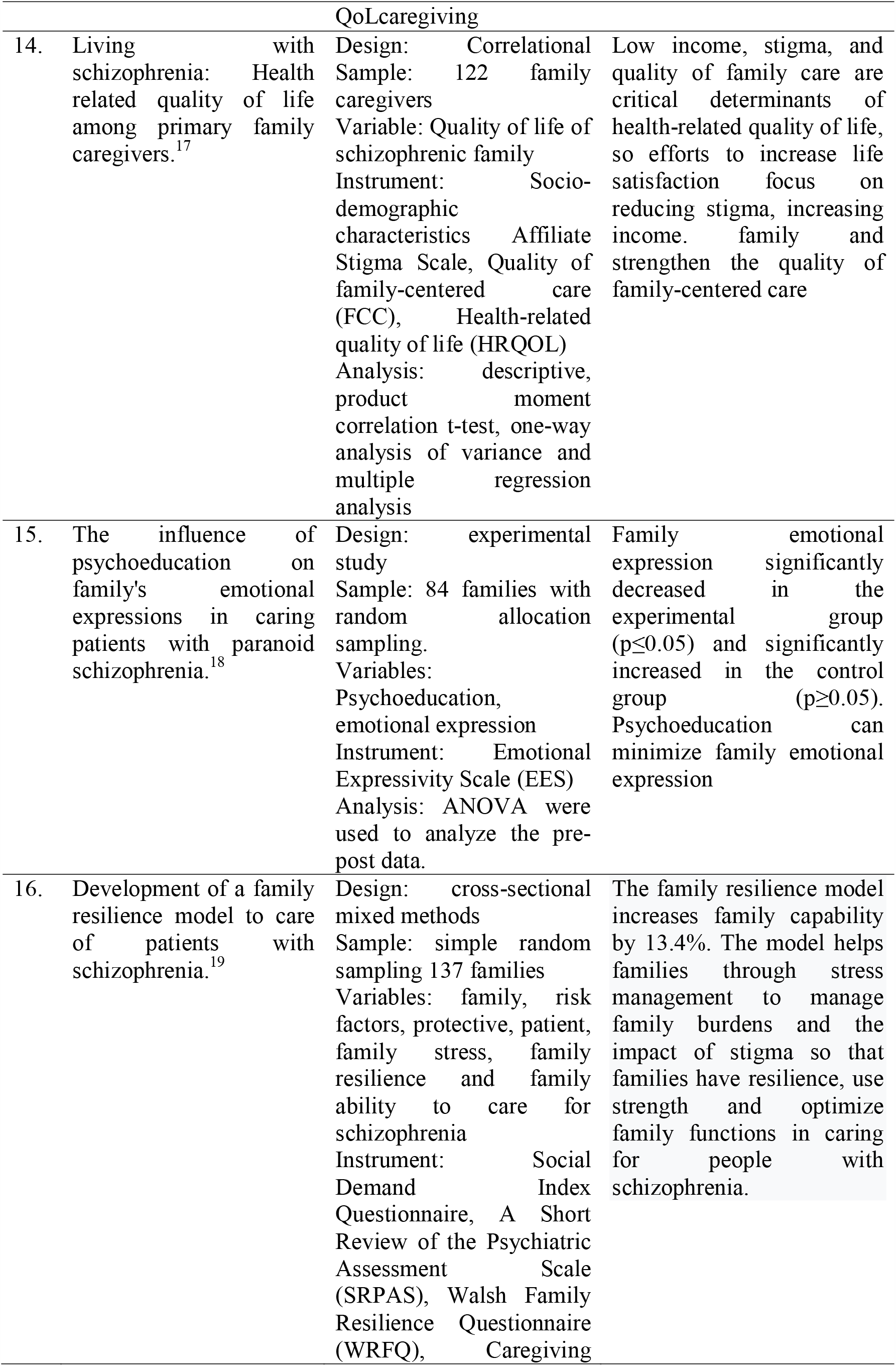
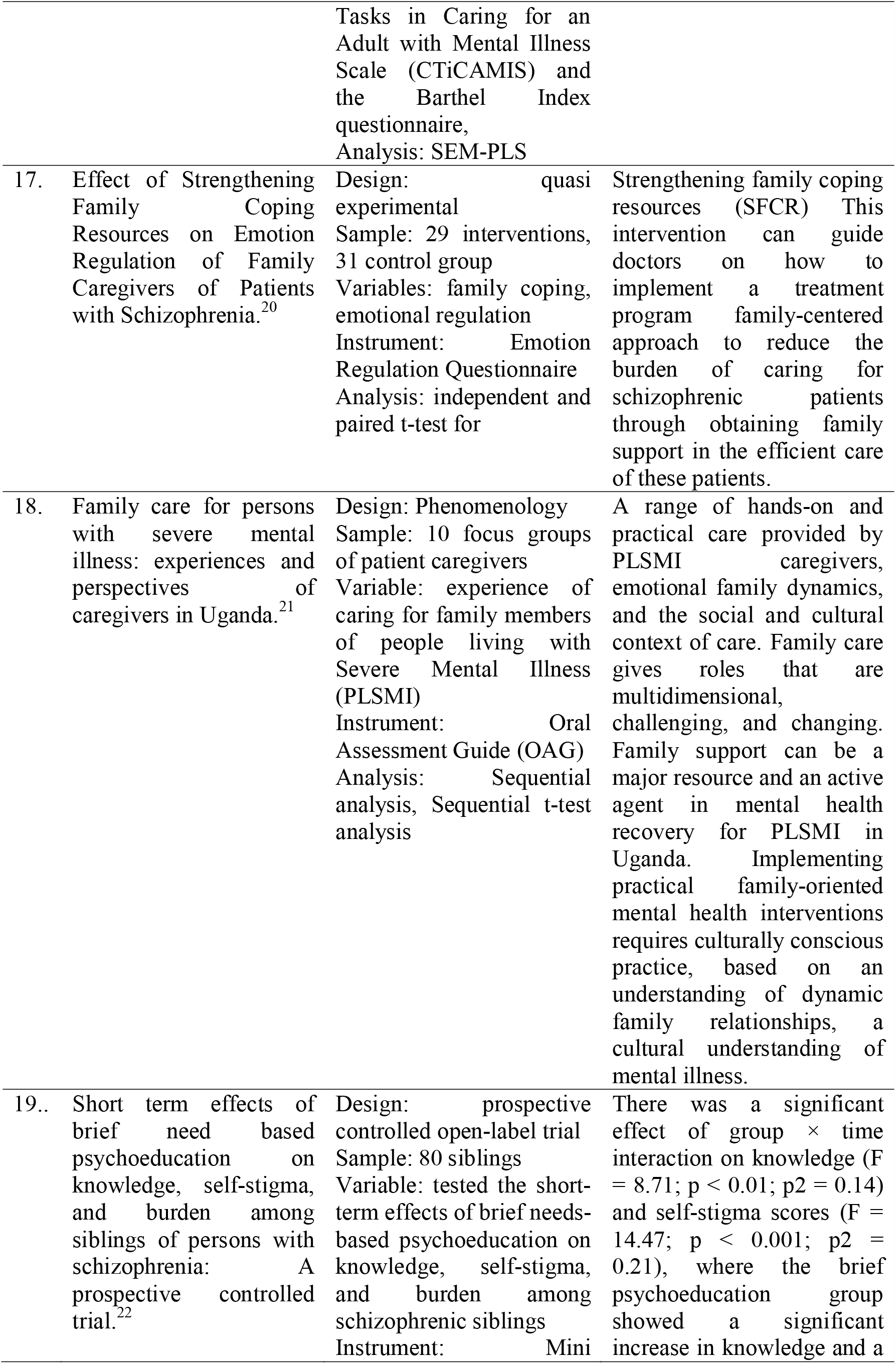
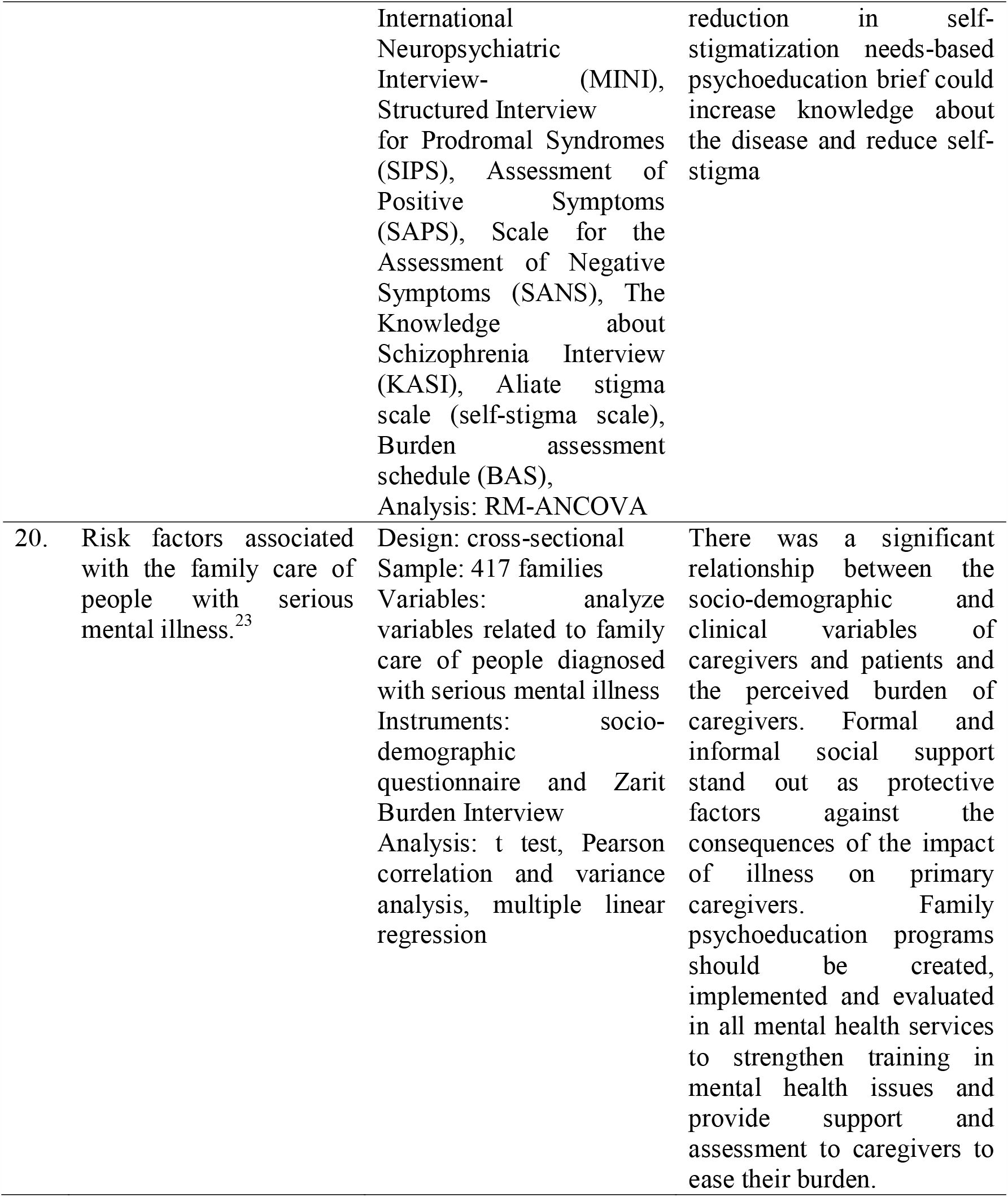

## DISCUSSION

### The Burden of Family Care

In many societies, family members are now the primary caregivers of mental health patients, taking on responsibilities traditionally under the overview of hospital and medical professionals. The impact of this shift on the family is high, has an emotional and economic burden.^15^ Families with schizophrenia experience the strongest stressors on the burden of care borne, in addition to stigma and the frequency of patient relapse.^3^ This is reinforced by the high household care burden for people living with schizophrenia in low-income settings.^16^ It was also found that caregiver of people with psychosis had worse mental health than non-caregivers. Single caregivers and those who spend more than 35 hours per week caring are found to be most at risk for poor mental health.^10^ Basically, the condition of schizophrenia has an impact on the burden on the family in caring for and it is proven that there is a significant relationship between socio-demographic and clinical variables of caregivers and patients and the burden felt by caregivers.^23^

The burden of treatment costs felt by the family is due to the length of recovery at the service provider and the frequency of recurrence, resulting in shorter re-admission times. The findings explain that low income, stigma, and quality of family care are critical determinants of health-related quality of life, so efforts to improve family life satisfaction are focused on reducing stigma, increasing family income and strengthening quality-centered care family. ^17^

### Family Psychoeducation

Mental health professionals can provide family support as a result of the burden of caring for schizophrenia by reducing the burden, family therapy, education and psychological support should be provided to families.^9^ This is reinforced by the finding that there is a need to provide support for caregivers by a mental health care delivery service system.^1^ Family psychoeducation can be an intervention that is considered to reduce the burden of family care, this is reinforced that Schizophrenic Patients who receive Family psychoeducation show a significant effect of treatment because they have a greater reduction in psychotic symptoms (PANSS), improvement in social functioning (SFQ), quality of life. life (QoL), and attitudes towards drugs (DAI).^11^ The psychosocial burden experienced by the family can also bring up emotional expressions that can be a barrier factor for care between patients and families, nursing interventions carried out to minimize emotional expressions have been widely studied and the majority recommend providing psychoeducation and the results are proven to reduce family emotional expressions.^18^

Mental health care professionals should involve all family members, including siblings, in the treatment plan for people with schizophrenia, and provide appropriate psychoeducation for all family members to reduce not only current parenting tensions and stress, but also anxiety about future care.^14^ The need for psychoeducation involving siblings in the management of schizophrenia is also reinforced by the finding that brief needs-based psychoeducation can increase knowledge about the disease and reduce self-stigma.^22^. Families will adapt and develop to respond to members who experience schizophrenia, family interventions that can be provided usually include psychoeducation, stress management, emotion, processing, assertive communication, cognitive reassessment, and structured problem solving proven to be effective in improving symptomatology, disease awareness, and levels of satisfaction. in patients and relatives.^15^

Components in psychoeducation in addition to family burden stress management also include family coping skills education in adapting to stressors in caring for families with schizophrenia, the results of the study explain that strengthening family coping resources (SFCR) This intervention can guide health professionals in implementing family-centered care programs to reduce stress. burden of caring for schizophrenic patients through family support in the efficient care of these patients.^20^. This is reinforced by the finding that there is a relationship between knowledge, perception, attitude, and family support and independence in treating hebephrenic schizophrenia.^7^ In addition, good support from health professionals and social services is needed to help family caregivers cope well with their stressors.^8^

Another approach used to reduce the burden experienced by families in caring for schizophrenia is to develop family resilience, which can increase the capability of families in caring for schizophrenia. The model helps families through stress management to reduce the burden of parenting and labeling sufferers. have resilience, use sources of strength and develop according to family functions to treat schizophrenia.^19^ The results of another study stated that strengthening family coping resources (SFCR) can guide service providers on how to implement family-centered care programs to reduce the burden of caring for schizophrenic patients through family support in efficient care.^20^

Care providers in the community can involve the family in a variety of ways, such as providing psychoeducation, supporting the family’s physical, emotional and social needs, and behavioral family assessment or family therapy.^24^ In addition, family psychoeducation programs should be created, implemented and evaluated in all mental health services to strengthen training in mental health issues and provide support and assessment to caregivers to ease their burden.^23^

## CONCLUSIONS

Family psychoeducation is an intervention of family support reducing the burden of caregivers taking care of schizophrenia. Mental health professionals need to develop and conduct family psychoeducation training so that it can be implemented in services.

## Data Availability

The data that support the findings of this study are available from the corresponding author.

